# Whole genome and transcriptome sequencing in neuromuscular disorders: a diagnostic and health economic analysis

**DOI:** 10.1101/2023.12.21.23300182

**Authors:** Ziying Yang, Xiaoru Yang, Yunmei Chen, Zhonghua Wang, Xiangke Fu, Lijie Song, Xunzhe Yang, Zhiyu Peng, Yi Dai

## Abstract

**Background:** A considerable number of patients suffering from neuromuscular disorders (NMD) are unable to receive an accurate diagnosis through initial genetic testing. It is imperative to develop a cost-effective diagnostic strategy that incorporates appropriate multi-omics techniques.

**Methods:** This study included 33 NMD patients with negative results from whole-exome sequencing (WES). Whole-genome sequencing (WGS) and RNA sequencing (RNA-seq) were performed concurrently to evaluate clinical utility. Additionally, eight diagnostic pathways were compared in terms of diagnostic rate, turnaround time, and cost.

**Results:** Our implementation of parallel WGS and RNA-seq testing successfully validated the clinical utility of this strategy in the cohort of 33 NMD patients initially yielding negative results from WES. The combined utilization of both methods resulted in an additional diagnosis for 42% (15/33) of the patients, with WGS contributing to 36% and RNA-seq contributing to 6% of the diagnoses. The Integration of alternative splicing results derived from RNA-seq data into variant filtering significantly reduced the number of rare intronic variants requiring interpretation and provided compelling evidence to support the classification of variant pathogenicity based on functional impact. Our comprehensive analysis, comparing eight different diagnostic pathways, revealed the cost-effectiveness of parallel WGS and RNA-seq testing as a diagnostic approach for patients. Moreover, the analysis of rare genomic findings within our cases showcased their potential to inform patient care, aid treatment decisions, and expand the range of NMD mutations in diagnosing rare NMD cases.

**Conclusion:** The integration of parallel WGS and RNA-seq testing represents a transformative diagnostic approach for NMD patients. The cost-effectiveness of this approach, coupled with its ability to improve diagnostic yield and interpretation efficiency, makes it a highly recommended strategy for clinical implementation to enhance the management and care of NMD patients.

## Introduction

Neuromuscular disorders (NMD) are rare diseases that involve muscles, motor neurons, peripheral nerves, or the neuromuscular junction. The clinical presentation and prognosis of NMD are highly variable, often leading to lifelong functional impairment and burden. Accurate diagnosis is crucial for appropriate monitoring and treatment.

Traditionally, the diagnosis of NMD relies on a combination of clinical and neurological examinations, electrophysiological studies of the peripheral nervous system, and histopathology of the muscle biopsy [1]. However, these clinical approaches have limitations when dealing with complex phenotypes or difficult differential diagnoses. The advent of massive parallel sequencing (MPS), notably the widely used whole exome sequencing (WES), has revolutionized our understanding of the genomic landscape of various NMD subtypes. MPS has emerged as a valuable tool for refining or altering NMD diagnoses [2, 3], providing prognostic information and family risk assessment [4], optimizing treatment options [5, 6], and identifying pharmacogenetically significant variants [5, 7]. Nonetheless, despite the advantages of MPS, approximately half of the patients still do not receive a molecular diagnosis after initial diagnostic workup [8–11].

Inconclusive cases can be partially attributed to challenges in variant detection and interpretation. WES has limitations in identifying coding and non-coding variants in genes encoding giant muscle proteins such as titin (TTN), nebulin (NEB), and the ryanodine receptor (RYR1). It also struggles with detecting repeat expansions in genes associated with hereditary ataxias [12]. Whole genome sequencing (WGS), which has the potential to detect all genomic variants, can provide an additional diagnostic yield of approximately 5% [13, 14] in cases where WES yields negative results. However, the main challenge lies in the prioritization and interpretation of variants for clinical application. In recent years, the integration of RNA sequencing (RNA-seq) into the diagnosis of genetic diseases has improved the diagnostic yield by 7.5%-35% [15, 16] in patients with negative WES results. RNA-seq offers valuable insights into the biological impact of candidate variants.

Additionally, proteomics and methylation analyses have shown promise in certain NMD disorders, such as Duchenne Muscular Dystrophy (DMD) [13, 14], Myasthenia Gravis [15], and Facio-Scapulo-Humeral Dystrophy (FSHD) [16]. These analyses provide proteomic and epigenetic signatures, enabling accurate diagnosis in some patients. However, relying solely on individual technologies may lead to misleading conclusions, and performing multiple tests simultaneously may not be cost-effective. Therefore, it is essential to explore the combination of different techniques and determine the optimal testing pathway. This approach will provide physicians with the most valid information for diagnosis while minimizing the waiting time and expenses for patients.

The diagnostic yield reported for each technique in patients with genetic disorders suggests that WGS could serve as an alternative to WES, while RNA-seq could effectively complement DNA-level testing. Based on this premise, we hypothesized that parallel WGS and RNA-seq testing would provide a more comprehensive and accurate diagnosis of NMD and offer higher cost-effectiveness. To investigate this hypothesis, we enrolled 33 NMD patients who had previously received negative results from WES. We conducted both WGS and RNA-seq on these individuals. The objectives of our study encompassed three aspects: (1) to validate the interpretive advantages and clinical utility of parallel WGS and RNA-seq testing; (2) to provide clinicians and patients with information to aid in selecting testing strategies by comparing various approaches, including routine testing method, in terms of diagnostic rate, turnaround time (TAT), and cost; (3) to analyze rare genomic findings in these cases and demonstrate how these findings can enhance patient care and treatment, as well as expand the NMD variant spectrum for the diagnosis of rare NMD.

## Materials and methods

### Study design

This study was approved by the Ethics Committee of Peking Union Medical College Hospital (NO. JS-3130), and written informed consent was obtained from all participants. We conducted a two-platform sequencing of the whole genome and whole transcriptome on a cohort of 33 NMD patients who had previously received negative results from WES testing. Positive results derived from these two tests will be subjected to family co-segregation analysis, while the absence of candidate variants will prompt the exploration of other multi-omics techniques (**Fig. 1**).

**Fig. 1.**
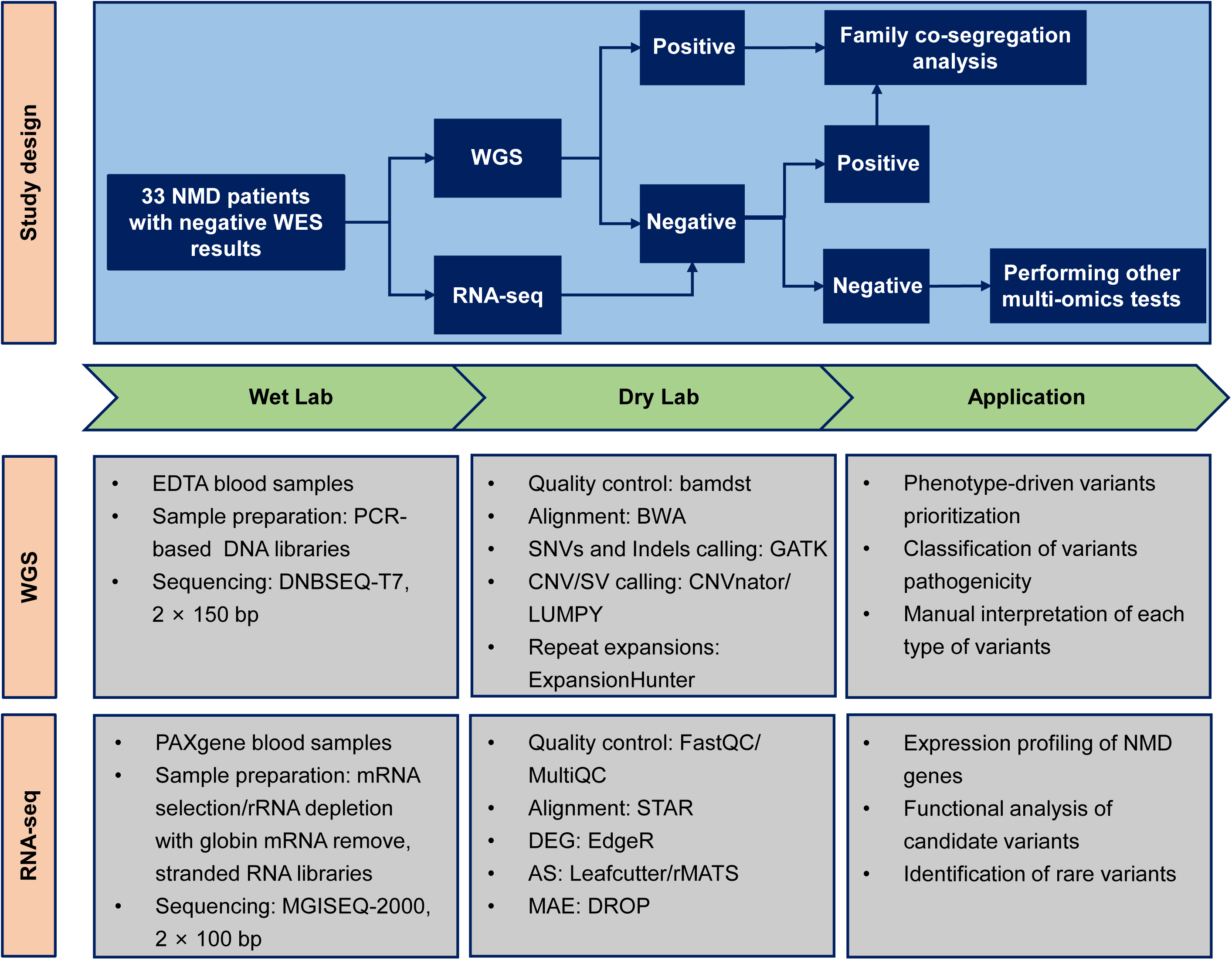
Study design. Parallel WGS and RNA-seq testing were conducted in the recruited NMD patients who had previously received negative WES results. It provides a comprehensive overview of the critical steps involved in the experimental process and the bioinformatics pipeline, including the software employed for both tests. NMD, Neuromuscular disorders; WGS, Whole genome sequencing; RNA-seq, RNA sequencing; WES, Whole exome sequencing.

### WGS

WGS libraries were prepared using the MGIEasy Enzymatic DNA Library Prep Kit v2.1 (MGI, Wuhan, China) following the manufacturer’s protocol, and then generated for DNA nanoballs. Short-read sequencing was performed on the DNBSEQ-T7 platform (MGI, Wuhan, China) with 150 bp paired-end reads to obtain 40 × mean coverage. The sequencing reads were aligned to the genome reference (hg19/GRCh37) and single nucleotide variants (SNVs), small insertions and deletions (Indels), copy number variations (CNVs), mitochondrial (MT) variants, structural variants (SVs), repeat expansions, and absence of heterozygosity (AOH) were analyzed using pipelines previously described [17].

### RNA-Seq

Total RNA was isolated using the PAXgene Blood RNA Kit (Qiagen, Chatsworth, CA, USA), following the manufacturer’s instructions. RNA samples with RNA integrity number (RIN) values ≥ 7.0 were selected for further RNA processing. For poly A+ RNA selection and the elimination of globin mRNA during library preparation, we utilized the QIAseq FastSelect-Globin Kit (Qiagen, Chatsworth, CA, USA) with 500 ng RNA as input for each sample. In the case of the two remaining RNA samples, both globin mRNA and rRNA were depleted using the Ribo-off Globin & rRNA Depletion Kit (Vazyme, Nanjing, China). For all 33 samples, stranded RNA libraries were prepared using the VAHTS Universal V8 RNA-seq Library Prep Kit (Vazyme, Nanjing, China).

Approximately 70 M reads (14 Gb raw data) per sample were generated using 100 bp paired-end reads on the MGISEQ-2000 platform (MGI, Wuhan, China). The software used in the bioinformatics pipeline is shown in **Fig. 1**. Sequence data quality was evaluated using FastQC (v0.11.9) and MultiQC (v1.11). The STAR (v2.7.9) aligner was used to align clean reads to the human genome (hg19/GRCh37). After alignment, the final BAM files were quantified using HTseq (v1.99.2) by Ensembl annotations (Gencode, GRCh37, release 19) and then normalized to TPM (transcripts per million mapped reads) values using RSEM (v1.3.1). EdgeR (v3.36.0) and DROP (v1.2.1) were used to detect differentially expressed genes (DEGs), and alternative splicing (AS) events were identified by using Leafcutter (v0.2.9) and rMATS (v4.1.2) with default parameters.

### Variant prioritization and classification

The candidate variants derived from the WGS analysis of an individual were subjected to stringent filtering criteria, which considered factors such as allele frequency, conservation, and their impact on protein sequence or mRNA splicing. Subsequently, we employed the Exomiser tool to prioritize these variants, utilizing the Human Phenotype Ontology (HPO) to standardize clinical symptoms and classify pathogenicity based on the guidelines established by the American College of Medical Genetics and Genomics (ACMG) [18]. Our analysis specifically focused on variants in NMD genes [19] and disease-causing genes present in the Online Mendelian Inheritance in Man (OMIM) database (**Table S1**). We manually categorized these variants in clinical practice as pathogenic, likely pathogenic, or of unknown significance (VUS). Finally, a multidisciplinary team comprising clinicians, genetic counselors, and geneticists conducted a thorough review of all the identified candidate causative variants.

### Variant confirmation and functional analysis

To validate the candidate variants for the probands and determine the carrier status of their family members, if applicable, we employed various techniques including Sanger sequencing, qPCR, and capillary electrophoresis. To investigate the impact of the deep-intronic variant on genes that are not expressed in blood, we conducted an in vitro minigene splicing assay. Briefly, we generated a mutant minigene construct using pcMINI and pcDNA3.1 vectors and confirmed the resulting recombinant plasmids (wild-type and mutant) through Sanger sequencing. Following this, we separately transfected the wild-type and mutant plasmids into MCF-7 and HEK293T cells. Subsequently, RNA extraction and RT-PCR were performed to analyze the splicing patterns.

## Results

### Study participants

We enrolled a cohort of 33 individuals diagnosed with NMD who had previously undergone WES without receiving a conclusive diagnosis. This cohort consisted of 10 males and 23 females, spanning an age range of 3 to 53 years, with a median age of 29 years. Among the cohort, there were 19 adults and 14 children. Serological testing was conducted on all patients, while muscle biopsies were performed on only 14 patients who exhibited specific indications, such as myopathic changes in electrophysiology or elevated serum creatine kinase levels [20]. Consequently, in clinical practice, when molecular assays need to be conducted on multiple platforms, obtaining blood samples is more convenient compared to muscle biopsies. Furthermore, electromyography testing was performed on 30 patients in our cohort (**Fig. 2a**).

**Fig. 2.**
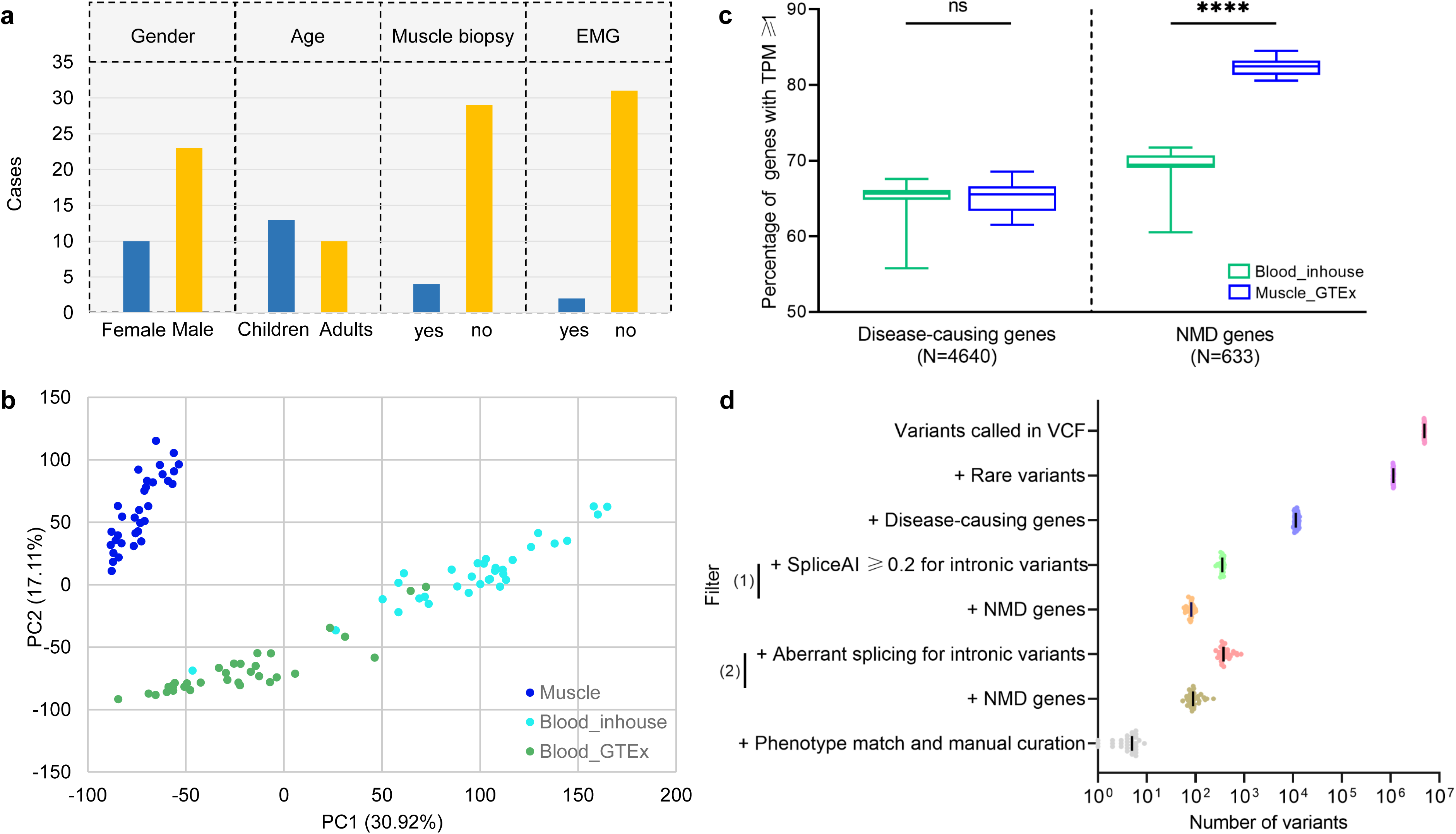
Patient characteristics and dual-platform sequencing features. **(a)** General information and the clinical test details of the patients. EMG, Electromyogram. **(b)** Principal component analysis plot of protein-coding gene expression (TPM) in RNA-seq data from our patients’ blood (blue-green), blood samples from the GTEx database (green), and muscle samples from the GTEx database (blue). TPM, Transcripts per million. GTEx, Genotypic Tissue Expression. **(c)** Percentage of disease-causing genes (N=4640) and NMD genes (N=633) expressed in blood data from our cohort (N=33) and muscle data from the GTEx database (N=33). The number of NMD genes expressed in muscle was significantly higher than in blood (p-value < 0.0001, Mann-Whitney test). However, it should be noted that more than two-thirds of the NMD genes were expressed in blood. **(d)** Variant filtering by integrating aberrant splicing results from RNA-seq data into WGS data. This approach substantially reduces the number of rare intronic variants requiring interpretation in disease-causing genes and NMD genes (TPM ≥ 1). VCF, Variant call format.

### Interpretation advantages and clinical utility of parallel testing of WGS and RNA-seq

Before interpreting the testing data, we first validated the reproducibility of the blood RNA-seq assay. We examined the consistency of protein-coding gene expression using RNA-seq data collected from the Genotypic Tissue Expression (GTEx) database.

Specifically, we analyzed 33 blood samples and 33 muscle samples, conducting principal component analysis (PCA) with our patients’ blood RNA-seq data. Our analysis yielded two distinct tissue clusters, with muscle samples exhibiting a higher degree of gene expression consistency in comparison to blood samples, as evidenced by the reduced variability observed in the former (**Fig. 2b**). Nevertheless, it is worth noting that blood RNA-seq still exhibited a high level of reproducibility, as the data from different sources remained closely aligned. Furthermore, we observed that the choice of RNA-selection methods could impact gene expression profiles, as indicated by the separation of the two samples prepared with rRNA deletion from those treated with mRNA-selection.

Subsequently, we proceeded to assess the expression levels of disease-causing genes (N=4640) and NMD genes (N=633) in whole blood. Our findings indicate that, on average, 65% of disease-causing genes and 69% of NMD genes were expressed (TPM ≥ 1) in our patients’ blood RNA-seq data. In comparison, the percentages of disease-causing genes and NMD genes expressed in muscle data from the GTEx database were 65% and 82%, respectively (p-value > 0.05, p-value < 0.0001, Mann Whitney test) (**Fig. 2c**). Consequently, these findings suggest that acquiring samples from the lesion site remains the preferable option for RNA-seq testing. However, blood RNA-seq provides an additional avenue for the clinical diagnosis of NMD patients, particularly when obtaining lesion tissues is challenging or invasive.

Subsequently, we evaluated the advantages of integrating RNA-seq results with WGS data for variant filtering and interpretation, as the task of identifying the candidate variant from the vast number of variants identified by WGS is complex. Nonetheless, the integration of RNA-seq results with WGS data offers an opportunity to enhance interpretation by providing functional insights into these variants, particularly in two significant ways. Firstly, by employing traditional filters such as population frequency and gene-disease association, the millions of variants identified in a single WGS dataset can be reduced to just over 10,000. Subsequently, by incorporating aberrant splicing (p-value ≤ 0.05) from the RNA-seq data as an additional filter, the number of rare intronic variants requiring interpretation on disease-causing genes (TPM ≥ 1) is significantly reduced, from a median of 11,201 to 144. This reduction in workload for interpretation, although not as substantial as the reduction achieved by implementing spliceAI score ≥ 0.2 [21] as a filter (93 variants, p-value < 0.01, Wilcoxon matched-pairs signed rank test), is still notable (**Fig. 2d**). A similar trend is observed when narrowing down the gene range to NMD genes, where fewer variants are filtered by aberrant splicing compared to spliceAI score (median number of 90 vs. 82, p-value < 0.01, Wilcoxon matched-pairs signed rank test). This discrepancy may arise from the failure of spliceAI to accurately predict the splicing effects of certain variants located deeper in introns, and the possibility of false-positive results in RNA-seq data. Secondly, the identification of aberrant splicing in RNA-seq data provides functional evidence within the framework of ACMG guidelines, such as PS3, which significantly impacts the pathogenicity classification of these variants.

Moreover, we assessed the clinical utility of parallel testing involving WGS and RNA-seq. In our cohort of 33 patients diagnosed with NMD, we detected pathogenic or likely pathogenic (P/LP) variants in 12 patients (36%), variants of uncertain significance (VUS) in 8 patients (24%), and no candidate variants in the remaining 13 patients (39%) through the interpretation of WGS data. Nevertheless, by integrating RNA-seq data into the analysis of WGS, we successfully upgraded VUS variants in 2 patients to P/LP variants, thereby increasing the total number of patients with P/LP variants to 14 (42%).

This transcriptome-guided genomic analysis provided an additional 10% diagnostic yield among the 21 patients who did not yield any P/LP variants through a single-platform WGS test (**Fig. 3**). Furthermore, we analyzed the diagnostic rates among patients with different phenotype subgroups. In patients with congenital myopathies, the diagnostic rate was 44% (4/9), while in those with muscular dystrophies, the rate was 40% (4/10). Both rates fall within the range of previously reported findings [22–24]. Notably, in congenital myopathies, which exhibit high genetic heterogeneity and nonspecific phenotypes, several candidate variants detected by WGS were not prioritized in the WES data. This may be due to their previous classification as VUS or because these variants did not match the patients’ phenotypes according to gene-disease associations. In contrast, muscular dystrophies display a relatively specific phenotype and a more limited genetic landscape.

**Fig. 3.**
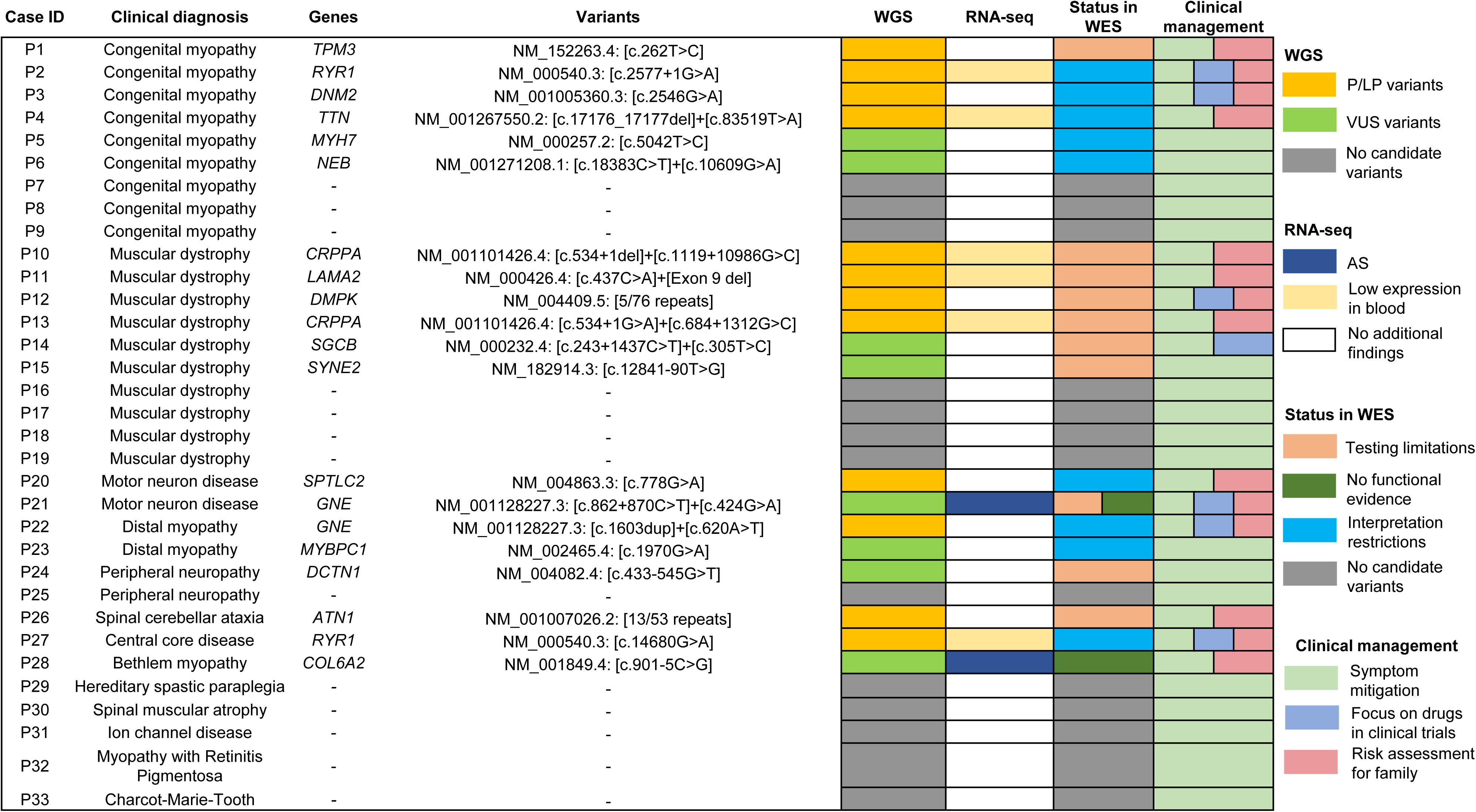
Clinical utility of parallel WGS and RNA-seq testing. A total of 14 patients (42%) were diagnosed with P/LP variants. WGS contributed to a diagnostic yield of 36% while the transcriptome-guided genomic analysis added 6% to the diagnostic yield.

Consequently, the candidate variants additionally detected by WGS are more likely to be the types of variants that WES struggles to identify, such as repeat expansions and small copy number variations specific to single exons, or variants located in non-coding or poorly captured regions that are undetectable by WES. However, despite these distinctions, the specificity of the clinical phenotype did not significantly impact the diagnostic rate through this dual-platform test (p > 0.05, Fisher’s exact test) across the groups of congenital myopathies, muscular dystrophies, and other diseases. This suggests that there is no requirement to select a specific phenotype for the clinical utilization of this assay. However, a more specific clinical phenotype may provide advantages in prioritizing candidate variants. Finally, among the 28 candidate variants identified through the dual-platform approach of WGS and RNA-seq, it was found that after excluding variants that could potentially be identified through WES reanalysis, the majority (60%) of the remaining 10 variants that were undetectable by conventional WES were intronic variants, followed by repeat expansions, which accounted for 20% of these variants.

### Health economic analysis of different diagnostic pathways

To develop an evidence-based clinical practice recommendation regarding the application of WES, WGS, and RNA-seq, we conducted a health economic analysis by evaluating the overall diagnostic rate, TAT, and cost of eight diagnostic pathways, which encompass various combinations of these three technologies (**Table 1**). The diagnostic pathways can be broadly categorized into two primary groups: sequential testing, where patients who tested negative in a previous round proceeded to another test, and concurrent testing. Given the substantial disparities in genomic sequencing costs (WES: US$800 (PerkinElmer laboratory)-US$936 (Centogene laboratory); WGS: US$1,628 (VCGS laboratory)-US$2,466 (Centogene laboratory) [25]; RNA-seq: US$390 (SeqCenter laboratory) (https://www.seqcenter.com/history)-US$1,930 (Emory NPRC Genomics Core)( https://enprc.emory.edu/nhp_genomics_core/rates/index.html)) and TAT (WES: 4 weeks (PerkinElmer laboratory)-10 weeks (Baylor genetics); WGS: 4 weeks (Centogene laboratory)-10 weeks (Baylor genetics); RNA-seq: 4 weeks-6 weeks (The St. Jude children’s research hospital) [26] across different laboratories, we opted to consider the lowest cost and shortest TAT among the available options for statistical analysis.

**Table 1.** Health economic analysis of 8 different testing modes.

| Testing modes | Parameters | WES<sup>a</sup> | WGS | RNA-seq | Total | Incremental | P-value<sup>d</sup> |
| --- | --- | --- | --- | --- | --- | --- | --- |
| **WES→(-)→WGS**<br>**→(-)→RNA-seq** | Diagnostic yield | 18%<br>(6/33) | 22% (6/27) | 10%<br>(2/21) | 42% (14/33) | 21% | / |
|  | TAT<sup>b</sup> | 4 weeks | 4 weeks | 4 weeks | Median TAT: 12 weeks | + 4 weeks | <0.0001 |
| | Cost | \$26,400<br>(\$800*33) | \$43,956<br>(\$1,628*27) | \$8,190<br>(\$390*21) | \$78,546 | \$41,616 | <0.0001 |
| **WES→(-)→RNA-seq** | Diagnostic yield | 18%<br>(6/33) | / | 3% (1/27) | 21% (7/33) | / | / |
|  | TAT | 4 weeks | / | 4 weeks | Median TAT: 8 weeks | / | / |
| | Cost | \$26,400<br>(\$800*33) | / | \$10,530<br>(\$390*27) | \$36,930 | / | / |
| **WGS→(-)→RNA-seq** | Diagnostic yield | / | 36% (12/33) | 10%<br>(2/21) | 42% (14/33) | 21% | / |
|  | TAT | / | 4 weeks | 4 weeks | Median TAT: 8 weeks | 0 | 0.0156 |
| | Cost | / | \$53,724<br>(\$1,628*33) | \$8,190<br>(\$390*21) | \$61,914 | \$24,984 | <0.0001 |
| **WES→(-)→WGS** | Diagnostic yield | 18%<br>(6/33) | 22% (6/27) | / | 36% (12/33) | 15% | / |
|  | TAT | 4 weeks | 4 weeks | / | Median TAT: 8 weeks | 0 | / |
| | Cost | \$26,400<br>(\$800*33) | \$43,956<br>(\$1,628*27) | / | \$70,356 | \$33,426 | <0.0001 |
| **WES+WGS+RNA-seq** | Diagnostic yield | 18%<br>(6/33) | 18% (6/33) | 6% (2/33) | 42% (14/33) | 21% | / |
|  | TAT | 4 weeks | 4 weeks | 4 weeks | Median TAT: 4 weeks | - 4 weeks | <0.0001 |
| | Cost | \$26,400<br>(\$800*33) | \$53,724<br>(\$1,628*33) | \$12,870<br>(\$390*33) | \$92,994 | \$56,064 | <0.0001 |
| **WES+RNA-seq** | Diagnostic yield | 18%<br>(6/33) | / | 3% (1/33) | 21% (7/33) | 0 | / |
|  | TAT | 4 weeks | / | 4 weeks | Median TAT: 4 weeks | - 4 weeks | <0.0001 |
| | Cost | \$26,400<br>(\$800*33) | / | \$12,870<br>(\$390*33) | \$39,270 | \$2,340 | 0.0313 |
| **WGS+RNA-seq** | Diagnostic yield | / | 36% (12/33) | 6% (2/33) | 42% (14/33) | 21% | / |
|  | TAT | / | 4 weeks | 4 weeks | Median TAT: 4 weeks | - 4 weeks | <0.0001 |
| | Cost | / | \$53,724<br>(\$1,628*33) | \$12,870<br>(\$390*33) | \$66,594 | \$29,664 | <0.0001 |
| **WGS** | Diagnostic yield | / | 36% (12/33) | / | 36% (12/33) | 15% | / |
|  | TAT | / | 4 weeks | / | Median TAT: 4 weeks | - 4 weeks | <0.0001 |
| | Cost | / | \$53,724<br>(\$1,628*33) | / | \$53,724 | \$16,794 | <0.0001 |

<sup>a</sup> It was assumed that the 6 cases previously undiagnosed due to interpretation restrictions could now be diagnosed using WES.
<sup>b</sup> It was assumed that there would be no interval between tests for the sequential diagnostic pathways. The statistical analysis considered the shortest TAT among laboratories: 4 weeks (PerkinElmer laboratory)- 10 weeks (Baylor genetics); WGS: 4 weeks (Centogene laboratory)- 10 weeks (Baylor genetics); RNA-seq: 4 weeks - 6 weeks (The St. Jude children's research hospital)
<sup>c</sup> The statistical analysis considered the lowest cost among laboratories: WES: US\$800 (PerkinElmer laboratory)-US\$936 (Centogene laboratory); WGS: US\$1,628 (VCGS laboratory)-US\$2,466 (Centogene laboratory); RNA-seq: US\$390 (SeqCenter laboratory)-US\$1,930 (Emory NPRC Genomics Core)
<sup>d</sup> P-values were calculated using the Wilcoxon matched-pairs signed rank test.
WGS, Whole genome sequencing; RNA-seq, RNA sequencing; WES, Whole exome sequencing; TAT, Turnaround time.

Additionally, we made an assumption that the six patients who had remained undiagnosed by WES due to restrictions in interpretation could potentially be identified through WES reanalysis.

Consistent with prior research findings, utilizing WGS as the initial genomic test has shown to yield higher diagnostic rates (36% vs. 18%) within the same TAT compared to when compared to WES, if the testing is solely conducted at the DNA level. Moreover, choosing WGS as the initial test is also proved to be a cost-effective option, as it achieves the same diagnostic outcomes at a lower cost than WGS following WES. The implementation of RNA-seq alongside genomic testing further enhances the diagnostic rate, although not as significantly as WGS. However, RNA-seq testing is less expensive, and the subsequent cost savings in healthcare expenses far outweigh the total diagnostic costs incurred. This makes the addition of RNA-seq a highly cost-effective strategy. In line with commonly recommended practices (RNA-seq after negative WES results) [27, 28], we selected this as our baseline model, and then calculated the incremental improvements in diagnostic rate, TAT, and cost for alternative diagnostic pathways. Our analysis showed that for sequential testing pathways, the diagnostic rate remained unaltered while the total cost decreased, and the TAT extended when compared with their corresponding concurrent testing pathways. However, it is worth noting that longer intervals between sequential tests may incur intangible healthcare costs. Additionally, the decreasing cost of sequencing is narrowing the cost gap between the two strategies. Moreover, timely diagnosis not only alleviates the stress experienced by families but also saves children from unnecessary tests and inappropriate treatments. Overall, conducting parallel testing of WGS and RNA-seq proves to be a cost-effective diagnostic pathway. In addition, from an economic perspective, the average annual healthcare cost for Duchenne Muscular Dystrophy beyond 5 years of age is USD 35,253 (https://everylifefoundation.org/burden-study), Based on this estimation, the annual healthcare cost for the 33 patients included in this study would amount to USD 1,163,349. Considering that the total diagnostic cost of USD 66,594 accounts for a mere 5.72% of this annual expenditure, it becomes evident that the cost of WGS and RNA-seq is relatively modest. This observation highlights the relatively minor financial burden imposed by WGS and RNA-seq, especially when compared to the lifelong healthcare expenses associated with rare disease management. However, it is crucial to acknowledge that the choice of a diagnostic pathway should not be fixed and should take into account factors such as a thorough clinical investigation, the urgency of testing, and the economic circumstances of the patient’s family.

### Genomic alterations of emerging biological and clinical relevance

The utilization of multi-omics approaches has revolutionized the diagnostic management of patients and significantly expanded our biomedical knowledge. In our study, the integration of RNA-seq data played a crucial role in the diagnosing two cases, providing invaluable insights. P28 presented with the phenotype of Bethlem myopathy and had previously been identified with a heterozygous c.901-5C>G variant in the *COL6A2* gene through WES data. Although this novel variant had been initially classified as VUS, its spliceAI score of 0.23 raised suspicions of its potential functional impact.

However, the analysis of RNA-seq data revealed exon 8 skipping, which could be attributed to this variant (**Fig. 4a**). As a result, an in-frame deletion occurred within the triple helical domain of the α2 (VI) chain. This deletion could hinder the proper assembly of α2 (VI) chains into dimers and tetramers, leading to a reduced production of collagen VI. Consequently, the patient’s diagnosis was confirmed, prompting a shift in clinical care towards comprehensive monitoring of complications and implementation of physical therapy. For instance, forced vital capacity measurements will be taken to prevent nocturnal hypoventilation resulting from disproportionate diaphragm weakness, and exercises aimed at joint stretching and muscle conditioning will be prescribed. If necessary, Achilles tendon release surgery can be considered to improve range of motion.

**Fig. 4.**
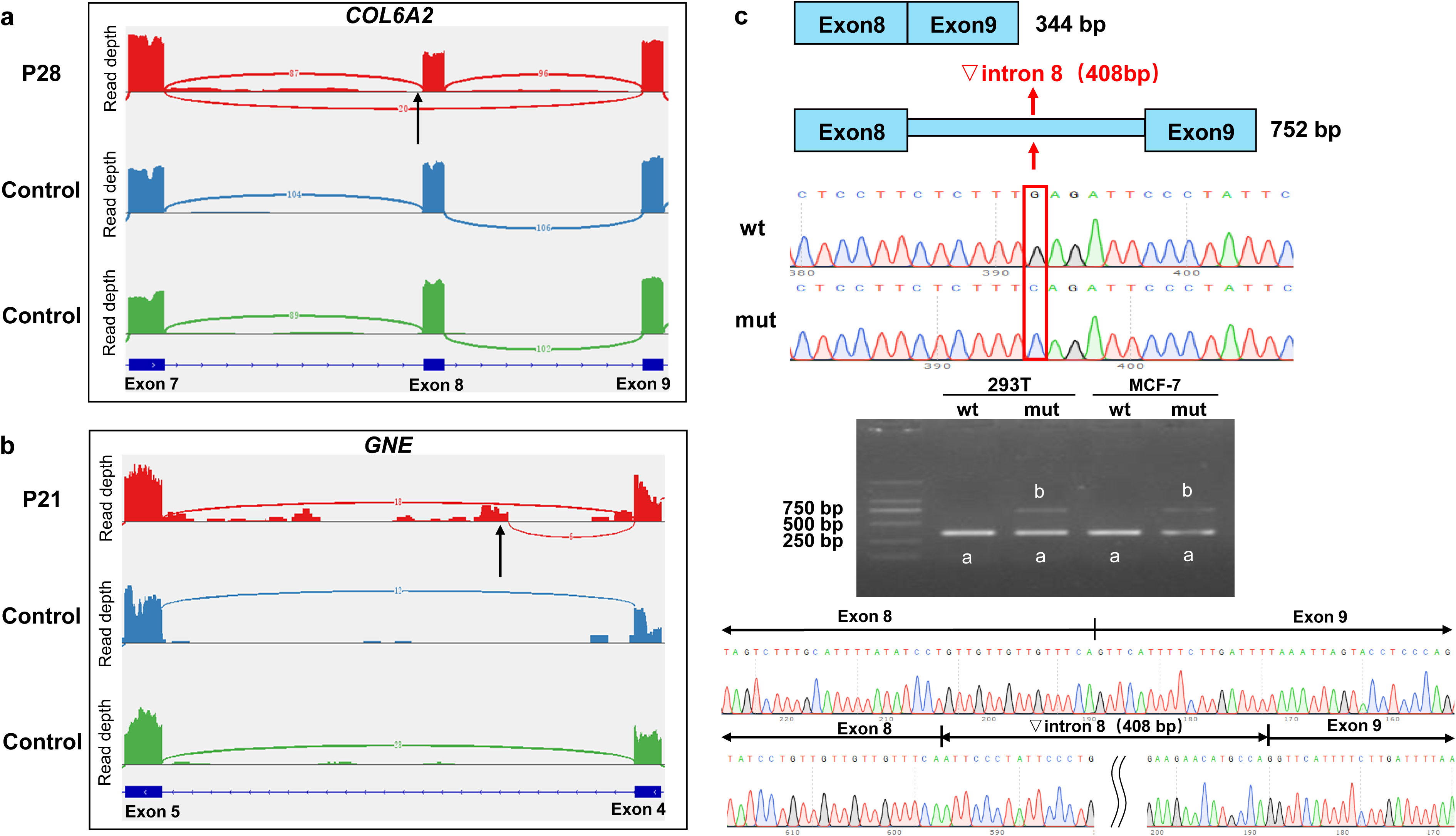
Rare genomic findings in cases. **(a)** P28 with Bethlem myopathy exhibited exon skipping of exon 8 due to a heterozygous c.901-5C>G variant on the *COL6A2* gene. The arrows point to the location where the variant occurred. **(b)** P21’s WGS data revealed compound heterozygous variants, c.424G>A and c.862+870C>T, on the *GNE* gene. The RNA-seq data indicated that the variant c.862+870C>T caused the retention of 723 bp in intron 4 of the *GNE* gene. **(c)** Analysis of the minigene splicing assay of variant c.1119+10986G>C on the *CRPPA* gene, identified in P10, demonstrated a retention of 408 bp in intron 8 through Sanger sequencing of reverse-transcription polymerase chain reaction products from transfected wild-type (wt) and mutant (mut) recombinant plasmids in MCF-7 and HEK293T cells.

In the case of P21, who presented with limb muscle weakness and difficulty in navigating stairs, WES data had initially identified a heterozygous c.424G>A variant in the *GNE* gene. However, due to the autosomal recessive inheritance pattern of the *GNE* gene, a definitive diagnosis could not be reached in the absence of the co-segregating variant. Nonetheless, WGS revealed an additional heterozygous c.862+870C>T variant in the deep intronic region of the *GNE* gene. This variant, with a spliceAI score of 0.71, was predicted to result in the retention of intronic region. Subsequent examination of blood RNA-seq data demonstrated the retention of 146 bp in intron 4 of the *GNE* gene caused by this variant. However, it is worth noting that the reads supporting splicing at the chr9:36235961 locus were found to be less than five after applying a filtering threshold (**Fig. 4b**). The combined utilization of WGS and RNA-seq allowed for the identification of another likely pathogenic variant in the *GNE* gene, ultimately leading to the patient’s diagnosis. The subsequent clinical management of this patient primarily focuses on providing symptomatic relief, including the use of assistive ambulatory devices (e.g., ankle-foot orthoses, canes, walkers, wheelchairs, or powerchairs) to address issues secondary to muscle weakness, as well as the use of adaptive devices to support fine motor function and activities of daily living. As ongoing clinical trials investigate drugs targeting the *GNE* gene [29, 30], it is essential to monitor research progress for potential future therapeutic applications.

Furthermore, the identification of pathogenic variants through dual-platform WGS and blood RNA-seq testing can offer valuable information for assessing the genetic risk of NMD in family members. In the case of P26, a diagnosis of pathogenic repeat expansion with over 48 CAG repeats on the *ATN1* gene was made. Notably, four individuals in his family exhibit hereditary ataxia, confirming their status as carriers of the pathogenic repeat expansion on the *ATN1* gene through capillary electrophoresis. This comprehensive testing approach enables accurate and comprehensive genetic risk assessment for family members, facilitating future genetic counseling.

### Complementary techniques enhance the elucidation of potential pathogenic variants

Given that some NMD genes are not expressed in blood, complementary techniques are required to assess the biological functional impact of candidate VUS variants identified through WGS. To address this, our study initiated an investigation into additional candidate variants within deep-intronic regions identified through WGS. Specifically, minigene assay was performed for P10 who carried a candidate intronic variant with a high spliceAI score but low expression of the corresponding gene in blood. P10 presented with mild muscular dystrophy, and WES analysis revealed a heterozygous c.534+1del variant in the *CRPPA* gene, which was absent from all population databases explored, including gnomAD and our in-house database containing data from over 9000 healthy individuals. An intriguing observation is that this variant is homozygous in the unaffected mother, which deviates from previously documented cases [31]. In those cases, bi-allelic null variants affecting the first five exons of the *CRPPA* gene were found to be associated with Walker-Warburg syndrome. Several potential mechanisms could account for this discrepancy. For instance, it is possible that the truncated protein retains partial activity, or there might be an incomplete nonsense-mediated decay process. In the case of our patient, WGS analysis revealed an additional c.1119+10986G>C variant on the gene, inherited from the healthy father. The minigene validation experiment indicated that the c.1119+10986G>C variant led to abnormal splicing within the mRNA, resulting in the retention of 408 bp of intron 8 at c.1119+10989_1119+11396 (**Fig. 4c**). This finding is consistent with the spliceAI prediction, which subsequently alters the reading frame and generates a premature termination codon, thereby producing a truncated protein comprising 417 amino acids.

## Discussion

In summary, we have successfully validated the clinical utility of parallel WGS and RNA-seq testing by implementing this strategy on a cohort of 33 patients with NMD who initially received negative results from WES. The combined utilization of both methods led to an additional diagnosis in 42% (15/33) of the patients, with WGS contributing to 36% and RNA-seq contributing to 6% of the diagnoses. By employing alternative splicing results derived from RNA-seq data as a filter, the number of rare intronic variants requiring interpretation was significantly reduced, thereby enhancing the efficiency of interpretation. Moreover, RNA-seq data provided compelling evidence of functional impact for the classification of variant pathogenicity. Furthermore, we conducted a comprehensive analysis comparing eight different diagnostic pathways, including the most recommended pathway, with respect to diagnostic rate, TAT, and cost. Our analysis revealed that parallel testing of WGS and RNA-seq proved to be a cost-effective diagnostic pathway for patients. Lastly, we analyzed the rare genomic findings observed in our cases and demonstrated their potential in informing patient care, facilitating treatment decisions, and expanding the spectrum of NMD mutations for the diagnosis of rare NMD cases.

Although the parallel testing of WGS and RNA-seq proved to be a powerful diagnostic tool in our study, there are still limitations to using clinically readily collected blood for RNA-seq testing. Firstly, approximately one-third of NMD genes either lack expression or exhibit low expression levels in the blood, resulting in reduced diagnostic yield as blood RNA-seq cannot provide evidence of biological impact for variants in these genes. Secondly, longer genes with multiple exons pose challenges in enrichment due to their extended mRNA lengths and high levels of blockage, despite being expressed in the blood. Thus, the effectiveness of blood RNA-seq in detecting these genes, particularly for *DMD* where intronic regions account for approximately 0.4% [32] of small pathogenic variants, is limited. It is likely that this data is underestimated, given the difficulties in conducting large-scale functional validation beyond muscle-specific RNA-seq data, as well as the invasive nature of muscle biopsies. To improve the diagnostic yield for patients and enhance the genomic profiles of these genes, it may be beneficial to increase the enrichment of these genes in blood RNA-seq through the use of target region capture techniques [33, 34], Additionally, exploring the use of other readily accessible tissue samples, such as cultured skin fibroblast cells [35] or renal epithelial cells [36, 37] could be considered. However, further evaluation of NMD-related gene expression in these samples is necessary.

Decisions regarding testing strategies remain crucial, as there is a trade-off between cost, diagnostic yield, and TAT. The optimal approach may vary depending on the specific use scenarios, such as prenatal diagnosis for high-risk pregnancies or diagnosis in neonatal or pediatric intensive care units (NICU/PICU), where prioritizing short TAT and high diagnostic rates is essential. While previous studies have primarily focused on comparative economic analyses of DNA testing techniques like WES, WES reanalysis, and WGS [25], few studies have incorporated RNA-seq into economic analyses though the value of multi-omics technologies, including RNA-seq, in clinical diagnosis of genetic disorders has been demonstrated. Our study contributes economic analyses for selecting pathways involving combinations of multi-omics technologies, providing valuable insights for the selection of genetic disease pathways. Nevertheless, it is important to acknowledge that our analysis is based on the lowest test prices and shortest TATs available among laboratories, which can significantly vary and lead to differing results. Additionally, factors such as additional expenses and time for hospitalization, counseling, and the impact of testing results on the quality of life for families, changes in management, and the benefits of accessing reproductive technologies should also be considered. Nonetheless, we anticipate that in the future, diagnostic rates and TAT will become increasingly critical as testing costs continue to decrease.

In our cohort, the parallel testing of WGS with RNA-seq yielded an additional diagnostic rate of approximately 42%, benefiting approximately half of the patients. However, the remaining half remained undiagnosed. For these individuals, alternative testing techniques can be considered based on the available genomic results. For instance, if a candidate VUS is identified but blood RNA-seq fails to provide evidence of biological impact, alternative approaches such as RT-PCR, minigene assay, or muscle tissue RNA-seq can be explored. In cases where no suspected variants are identified by genomic testing, long-read sequencing can be employed to screen for structural variants [38, 39], repeat expansions [40], and other limited types of variants not easily detected by short-read sequencing. Alternatively, it can be detected using multi-omics techniques such as proteomics analysis and methylation analysis. It is important to note that these tests are currently expensive and not yet widely available in clinical practice. We conducted long-read WGS in a patient diagnosed with Becker Muscular Dystrophy, but unfortunately, no candidate variants were identified. Furthermore, patients with a clear clinical diagnosis but no molecular diagnosis despite multiple testing techniques require ongoing attention. This may involve data reanalysis or the exploration of novel causative genes to address the diagnostic challenges.

The development of multi-omics technology has brought both benefits and challenges to the diagnosis of patients with genetic diseases. The value of each testing technology needs to be assessed through large-scale clinical applications, and the selection of the most optimal testing pathway to achieve maximum benefits at the lowest cost remains a challenge for healthcare providers and geneticists. In the future, larger cohorts and more comprehensive evaluations will be necessary for health economic analyses.

## Statements and Declarations

### Authors’ contributions

Y.D. designed the research. Z.Y. wrote the first draft of the article. X.Y. performed the experiments. Y.C. and Z.W. performed data analysis. X.F. performed the statistical analysis and XZ.Y. recruited patients and collected specimens. L.S. and Z.P. contributed to drafting and revising the manuscript. All authors reviewed the manuscript and approved the submitted version.

### Data Availability

The data that support the findings of this study have been deposited into the CNGB Sequence Archive (CNSA) [41] of China National GeneBank DataBase (CNGBdb) [42] with accession number CNP0004985. The datasets for this article are not publicly available due to privacy or ethical restrictions. Requests to access the datasets should be directed to the corresponding author.

### Ethics approval

Written informed consent was obtained from all the participants. This study was approved by the Ethics Committee of Peking Union Medical College Hospital (NO. JS-3130) and was performed by the Declaration of Helsinki.

## Acknowledgments

We thank all the blood donors for their invaluable contribution to this study.

## References

1. Yubero, D., et al., The Increasing Impact of Translational Research in the Molecular Diagnostics of Neuromuscular Diseases. Int J Mol Sci, 2021. 22(8).

2. Ziats, M.N., et al., Genotype-phenotype analysis of 523 patients by genetics evaluation and clinical exome sequencing. Pediatr Res, 2020. 87(4): p. 735–739.

3. Beecroft, S.J., et al., Targeted gene panel use in 2249 neuromuscular patients: the Australasian referral center experience. Ann Clin Transl Neurol, 2020. 7(3): p. 353–362.

4. Waldrop, M.A., et al., Diagnostic Utility of Whole Exome Sequencing in the Neuromuscular Clinic. Neuropediatrics, 2019. 50(2): p. 96–102.

5. Chikkannaiah, M. and I. Reyes, New diagnostic and therapeutic modalities in neuromuscular disorders in children. Curr Probl Pediatr Adolesc Health Care, 2021. 51(7): p. 101033.

6. Beecroft, S.J., et al., The Impact of Next-Generation Sequencing on the Diagnosis, Treatment, and Prevention of Hereditary Neuromuscular Disorders. Mol Diagn Ther, 2020. 24(6): p. 641–652.

7. Thompson, R., et al., Advances in the diagnosis of inherited neuromuscular diseases and implications for therapy development. Lancet Neurol, 2020. 19(6): p. 522–532.

8. Herman, I., et al., Clinical exome sequencing in the diagnosis of pediatric neuromuscular disease. Muscle Nerve, 2021. 63(3): p. 304–310.

9. Westra, D., et al., Panel-Based Exome Sequencing for Neuromuscular Disorders as a Diagnostic Service. J Neuromuscul Dis, 2019. 6(2): p. 241–258.

10. Chen, P.S., et al., Diagnostic Challenges of Neuromuscular Disorders after Whole Exome Sequencing. J Neuromuscul Dis, 2023.

11. Al Sultani, H., K. Hafeez, and A. Shaibani, Diagnostic Outcome of Genetic Testing for Neuromuscular Disorders in a Tertiary Center. J Clin Neuromuscul Dis, 2022. 24(1): p. 1–6.

12. Stranneheim, H., et al., Integration of whole genome sequencing into a healthcare setting: high diagnostic rates across multiple clinical entities in 3219 rare disease patients. Genome Med, 2021. 13(1): p. 40.

13. Schreyer, L., et al., The discovery of the DNA methylation episignature for Duchenne muscular dystrophy. Neuromuscul Disord, 2023. 33(1): p. 5–14.

14. Roos, A., et al., Intersection of Proteomics and Genomics to “Solve the Unsolved” in Rare Disorders such as Neurodegenerative and Neuromuscular Diseases. Proteomics Clin Appl, 2018. 12(2).

15. Lin, J., et al., Epigenome-wide DNA methylation analysis of myasthenia gravis. FEBS Open Bio, 2023. 13(7): p. 1375–1389.

16. Caputo, V., et al., D4Z4 Methylation Levels Combined with a Machine Learning Pipeline Highlight Single CpG Sites as Discriminating Biomarkers for FSHD Patients. Cells, 2022. 11(24).

17. Yang, Z., et al., Test development, optimization and validation of a WGS pipeline for genetic disorders. BMC Med Genomics, 2023. 16(1): p. 74.

18. Richards, S., et al., Standards and guidelines for the interpretation of sequence variants: a joint consensus recommendation of the American College of Medical Genetics and Genomics and the Association for Molecular Pathology. Genet Med, 2015. 17(5): p. 405–24.

19. Cohen, E., et al., The 2022 version of the gene table of neuromuscular disorders (nuclear genome). Neuromuscul Disord, 2021. 31(12): p. 1313–1357.

20. Walters, J. and A. Baborie, Muscle biopsy: what and why and when? Pract Neurol, 2020. 20(5): p. 385–395.

21. Jaganathan, K., et al., Predicting Splicing from Primary Sequence with Deep Learning. Cell, 2019. 176(3): p. 535–548 e24.

22. Cavdarli, B., et al., High diagnostic yield of targeted next-generation sequencing panel as a first-tier molecular test for the patients with myopathy or muscular dystrophy. Ann Hum Genet, 2023. 87(3): p. 104–114.

23. Vill, K., et al., Early-Onset Myopathies: Clinical Findings, Prevalence of Subgroups and Diagnostic Approach in a Single Neuromuscular Referral Center in Germany. J Neuromuscul Dis, 2017. 4(4): p. 315–325.

24. Sanga, S., et al., Whole-exome analyses of congenital muscular dystrophy and congenital myopathy patients from India reveal a wide spectrum of known and novel mutations. Eur J Neurol, 2021. 28(3): p. 992–1003.

25. Ewans, L.J., et al., Whole exome and genome sequencing in mendelian disorders: a diagnostic and health economic analysis. Eur J Hum Genet, 2022. 30(10): p. 1121–1131.

26. Nurchis, M.C., et al., Incremental net benefit of whole genome sequencing for newborns and children with suspected genetic disorders: Systematic review and meta-analysis of cost-effectiveness evidence. Health Policy, 2022. 126(4): p. 337–345.

27. Yepez, V.A., et al., Clinical implementation of RNA sequencing for Mendelian disease diagnostics. Genome Med, 2022. 14(1): p. 38.

28. Hong, S.E., et al., Transcriptome-based variant calling and aberrant mRNA discovery enhance diagnostic efficiency for neuromuscular diseases. J Med Genet, 2022. 59(11): p. 1075–1081.

29. Suzuki, N., et al., Phase II/III Study of Aceneuramic Acid Administration for GNE Myopathy in Japan. J Neuromuscul Dis, 2023. 10(4): p. 555–566.

30. Carrillo, N., et al., Safety and efficacy of N-acetylmannosamine (ManNAc) in patients with GNE myopathy: an open-label phase 2 study. Genet Med, 2021. 23(11): p. 2067–2075.

31. Cirak, S., et al., ISPD gene mutations are a common cause of congenital and limb-girdle muscular dystrophies. Brain, 2013. 136(Pt 1): p. 269–81.

32. 32. Tong, Y.R., et al., A Comprehensive Analysis of 2013 Dystrophinopathies in China: A Report From National Rare Disease Center. Front Neurol, 2020. 11: p. 572006.

33. Bouge, A.L., et al., Targeted RNA-Seq profiling of splicing pattern in the DMD gene: exons are mostly constitutively spliced in human skeletal muscle. Sci Rep, 2017. 7: p. 39094.

34. Okubo, M., et al., RNA-seq analysis, targeted long-read sequencing and in silico prediction to unravel pathogenic intronic events and complicated splicing abnormalities in dystrophinopathy. Hum Genet, 2023. 142(1): p. 59–71.

35. Gonorazky, H.D., et al., Expanding the Boundaries of RNA Sequencing as a Diagnostic Tool for Rare Mendelian Disease. Am J Hum Genet, 2019. 104(5): p. 1007.

36. Maddirevula, S., et al., Analysis of transcript-deleterious variants in Mendelian disorders: implications for RNA-based diagnostics. Genome Biol, 2020. 21(1): p. 145.

37. Lee, H., et al., Diagnostic utility of transcriptome sequencing for rare Mendelian diseases. Genet Med, 2020. 22(3): p. 490–499.

38. Bruels, C.C., et al., Diagnostic capabilities of nanopore long-read sequencing in muscular dystrophy. Ann Clin Transl Neurol, 2022. 9(8): p. 1302–1309.

39. Miyatake, S., et al., Rapid and comprehensive diagnostic method for repeat expansion diseases using nanopore sequencing. NPJ Genom Med, 2022. 7(1): p. 62.

40. Ibanez, K., et al., Whole genome sequencing for the diagnosis of neurological repeat expansion disorders in the UK: a retrospective diagnostic accuracy and prospective clinical validation study. Lancet Neurol, 2022. 21(3): p. 234–245.

41. Guo, X., et al., CNSA: a data repository for archiving omics data. Database (Oxford), 2020. 2020.

42. Chen, F.Z., et al., CNGBdb: China National GeneBank DataBase. Yi Chuan, 2020. 42(8): p. 799–809.

